# Improved COVID-19 testing by extraction free SARS-Cov-2 RT-PCR

**DOI:** 10.1101/2020.08.10.20171512

**Authors:** Khelil Mohamed Mokhtar

**Affiliations:** Pasteur institute of Algeria, department of immunology, Annex of M’sila, Algeria

**Keywords:** SARS-CoV-2, COVID 19, RT-PCR, heat-treatment, nasopharyngeal swab samples

## Abstract

The RNA extraction is an important checkpoint for the detection of SARS-CoV-2 in swab samples, but it is a major barrier to available and rapid COVID-19 testing. In this study, we validated the extraction-free RT-qPCR method by heat-treatment as an accurate option to nucleic acid purification in Algerian population.

---

Dear editor:

The new emergence of the novel human coronavirus in January 2019 in Wuhan City (China), rapidly evolved into a global pandemic. The virus was confirmed to have spread to Algeria in February 2020, which put notable pressure on public and private health laboratories as they attempt to keep up with demands for SARSCoV-2 testing despite shortage of reagents (1). Currently, the widely used protocol for SARS-CoV-2 detection is RT-qPCR assay preceded by purification of viral RNA from patient sample, typically from nasopharyngeal (NP) swab as described by CDC and WHO (2-4). However, nucleic acid purification step is not only laborious and time-consuming, but the additional steps requiring manual handling can result in experimental errors, especially false positive results due to specimen-to-specimen carryover (5). To address this issue, recent attempts have been made to circumvent RNA extraction in COVID-19 testing by performing RT-qPCR directly on heat-inactivated subject samples (65°C for 30 min or 95°C for 10min) or directly loading patient swab medium into RT-PCR reaction mix. Using heat-inactivation approach the sensitivity ranged from 92 to 96% and specificity from 93 to 100% (6). Here, we tested the direct method of SARS-CoV-2 RT-qPCR on heat-treated (Hit-RT-PCR) nasopharyngeal swab samples and compared the results with RNA-extraction based RT-PCR results.

This study was conducted at the clinical laboratory of INSTITUT PASTEUR OF M’SILA, ALGERIA. Clinical samples (nasopharyngeal swabs) from patients with high likelihood for COVID-19 were collected by medical infectiologists and deposited in viral transport medium at different healthcare institution of the city of M’sila. Arrived to the laboratory, samples were stored at -20°C until extracted and tested within 72h. For routine analysis, RNA was extracted from 140 μL of NP samples using the QIAamp Viral RNA Mini kit. Reverse transcription and quantitative PCR were performed using the Biogerm® novel Coronavirus (2019-nCoV) nucleic acid kit following the manufacturer instructions: Total reactions of 25μl were obtained by mixing 20μl of master mix (primers and probe mix: ORF1ab, N and RNase P genes) and 5 μl of clinical sample to fill the reaction. The thermal cycling steps were: Stage1: 50°C for 10 min, stage2: 95°C for 5 min, stage3: 95°C for 10 sec, 55°C for 40 sec, 40 cycles. The RT-qPCR was performed on a Rotor-Gen Q real time PCR machine (Qiagen®) using the Rotor-Gen Software v2.3.

We initially aimed to validate heat-treatment method to get an accurate view of its performance in a real world clinical diagnostic setting. We blindly heated a panel of aliquots from 60 NP samples representing intermediate (CT of 20 - 30) and low (CT of more than 30) viral RNA loads by direct RT-qPCR. The SARS-CoV-2 Ct levels (ORF1ab and N) in these samples were previously determined by RT-qPCR that included RNA extraction (Ct cutoff ≤38).

NP swab samples were thermally treated in water bath at 65°C for 30 min.Samples were then placed in room temperature for 15 min, vortexed for 10 seconds and 5 μl of the supernatant was directly loaded into RT-qPCR reaction. Comparably, aliquots from 161 NP samples were subjected to heat-treatment but with increasing heating time to 60 min.

An agreement analysis (positive and negative percent agreement) were applied between diagnostic results of our experiment and results obtained by the conventional SARS-CoV-2 testing protocol. Diagnostic results were considered as categorical variables (1 for the presence of SARS cov2 infection and 0 for the absence of infection). All statistical analysis were performed using R version 3.6.0 (R Core Team, 2014) (7). In this work we used anonymized material from samples that had been collected for clinical diagnostics of SARS-CoV-2.

We found a weak agreement when NP samples were heated for 30min (PPA: 58%, 95%CI: 45 to 69%).But, the agreement increased (PPA: 78%, 95%CI: 70 to 84%) when we increased the heating time to (60 min). We also found a substantial agreement between N gene results of extracted and heat-inactivated samples (overall agreement 78%, 95%CI 70 to 83%) but a weak agreement for ORF1ab gene (overall agreement 45%, 95%CI 37 to 52%). Ct values of N gene for hid-RT-qPCR samples were higher than for RNA eluates of the same samples (mean difference =1.9 Ct). Surprisingly, three samples were identified as COVID-19 positive by 60 min heat-inactivation RT-qPCR (one sample positif for N and ORF1ab and two for only N) but were negative on extracted RNA (Table.2, sample 17, 19 and 84). Supplementary data represented by Heatmap1 and 2 shows the full results of this experiment while Table 3 provides a summary.

**Table 3.** Detection sensitivity of Hic-RT-qPCR for 30 min versus Hic-RT-qPCR for 60 min on NP samples containing a range of SARS-CoV-2 viral RNA loads.

| **Viral RNA load**<br>**( Ct)** | **Heat-inactivation time** |  |
| --- | --- | --- |
|  | ***30 min*** | ***60 min*** |
| 20 - 30 | 26/34 (76%) | 74/74 (100%) |
| >30 | 9/26 (34%) | 38/70 (54%) |
| **Total** | **35/60 (58%)** | **112/144 (78%)** |

Clinical laboratories of the developing world are overwhelmed with COVID-19 testing demands. As a means to validate heat-treatment RT-PCR method in our clinical laboratory, we have shown that prior heating at 65°C for 30 min was less accurate compared to prior heating at 65°C for 60 min. Our observation were not corroborated by previous results which showed that prior heating at 65°C for 30 min was adequate to correctly identify 92 to 96% of screened samples. This could be explained by difference in the composition of viral transport medium used (Inhibitory agents from the swab and medium may inhibit RT-qPCR) or a mutations in the Algerian strain of SARS-cov2, rendering the virus more resistant to heat inactivation. Our improved protocol correctly identified 100% of clinical samples with viral load between (20 and 30 Ct). The only samples missed were those among lower Ct range (Ct> 30). Of the 2065 cases with a positive diagnosis at “Institut Pasteur of M’sila” by our clinical laboratory at the time of writing, only 27% would fall in this low Ct range, which demonstrate that our improved protocol will accurately detect the majority of COVID-19 patients. Evidence that analytical sensitivity of hid-RT-qPCR was inferior (higher Ct values) compared to extraction-based RT-qPCR is that heating for long time may degrade RNA in presence of metal ions and/or RNases and that more RNA was loaded for eluates compared to Hic-RT-PCR. Furthermore, the higher performance of primers and probes targeting short amplicon (N, 110 bp) confirmed previous reports. Hence, short amplicons targets may be more suitable for Hic-RT-qPCR protocol.

A surprising finding was that hic-RT-PCR identified three samples as COVID-19 positive while they had been identified as COVID-19 negative by conventional protocol. The Ct values of hic-RT-PCR samples were high (> 30) suggesting one possible explanation of this phenomenon: NP samples may had very low viral RNA load that was below the limit of detection - i.e the lowest concentration level with a detection rate of 95% for positive results- of the RT-PCR kit (1000 copies/ml) (9). So, negative results in patients with typical symptoms of COVID-19 may become detectable by repeating the test. Unfortunately, we were unable to confirmed COVID-19 positivity by collection of a new swab samples.

In summary we have shown that RT-qPCR testing for SARS-CoV-2 infection could be achieved through heat-inactivation protocol (65°C for 60 min) without the use of RNA extraction kits. Our findings suggest that heat-inactivation protocol may be useful for massive SARS-CoV-2 screening after being validated for each region. Previous reports suggest that initial negative result by Hic-RT-PCR should be repeated by RNA extraction for symptomatic patients, healthcare personnel, and others with a high likelihood of suspicion (8). However, based on recent evidence showing the oddity of SARS-CoV-2 that can be cultured in respiratory samples 9 days after symptom onset, notably in patients with mild disease, it appears that re-testing in such patients may not be necessary (10). Such a strategy would drastically reduce the need for RNA extraction for a substantial portion of future COVID-19 tests.

## Data Availability

we have provided a supplementary material which include some experimental data

## Acknowledgements

We are grateful to Mekki Oussama who participated in sample handling, organization and performed experiments.

